# Severe *GBA1* variants drive the GBA-PD clinical phenotype: implications for counselling and clinical trials

**DOI:** 10.1101/2024.12.09.24318560

**Authors:** Elisa Menozzi, Sara Lucas Del Pozo, Jane Macnaughtan, Roxana Mezabrovschi, Sofia Koletsi, Pierfrancesco Mitrotti, Luca Gallo, Rosaria Calabrese, Marco Toffoli, Nadine Loefflad, Franco Valzania, Francesco Cavallieri, Valentina Fioravanti, Selen Yalkic, Naomi Limbachiya, Fabio Blandini, Micol Avenali, Anthony HV Schapira

**Affiliations:** Department of Clinical and Movement Neurosciences, Queen Square Institute of Neurology, University College London (UCL), London, United Kingdom; Aligning Science Across Parkinson’s (ASAP) Collaborative Research Network, Chevy Chase, MD, 20815; Liver Failure Group, Institute for Liver and Digestive Health, University College London, London, UK; Department of Brain and Behavioral Sciences, University of Pavia, Pavia, Italy; IRCCS Mondino Foundation, Pavia, Italy; Neurology Unit, Neuromotor and Rehabilitation Department, Azienda USL-IRCCS di Reggio Emilia, Reggio Emilia, Italy; Ca’ Granda IRCCS Foundation, Ospedale Maggiore Policlinico, Milan, Italy

## Abstract

**Background:** Variants in the *GBA1* gene are the commonest genetic risk factor for Parkinson disease (PD). Genotype-phenotype correlations exist but with conflicting data, particularly in the cognitive domain.

**Objectives:** Comparing clinical phenotypes in a multicentre, international cohort incorporating GBA-PD and idiopathic PD (iPD) patients.

**Methods:** Patients underwent a comprehensive assessment of motor and non-motor functions. Two-group (GBA-PD vs iPD) and multiple-group comparisons (iPD, risk, mild, and severe variant GBA-PD) were performed.

**Results:** Three hundred eleven PD patients were recruited: 183 iPD, 39 severe GBA-PD, 24 mild GBA-PD, 55 risk GBA-PD, and 10 patients carrying variants of unknown significance. Groups were matched for sex, education, disease duration and medications. Mild and severe GBA-PD were younger and developed PD earlier. Severe GBA-PD had worse depression, cognitive impairment, hyposmia, and motor complications.

**Conclusions:** Only severe variant GBA-PD have a distinctive, more severe clinical profile.

## Introduction

Variants in the *GBA1* gene are highly prevalent in the Parkinson disease (PD) population (~10-15%).^1^ In recent years, the increased recourse to *GBA1* genotyping in PD,^2^ the different response of patients carrying *GBA1* variants (GBA-PD) to advanced treatments for PD (i.e., deep brain stimulation-DBS),^3^ and the growing number of clinical trials testing *GBA1*-targeted therapies,^4^ have created the need for standardized guidelines for genetic counselling, management and selection of patients for clinical trials.^1^

Major challenges in manifest GBA-PD include the differences in clinical severity and progression according to *GBA1* variant type.^4^ Several case-control studies have reported that PD patients carrying severe *GBA1* variants (e.g., p.L483P) compared to carriers of mild (e.g., pN409S) or risk (e.g., p.E365K) variants, present with a more severe clinical phenotype characterised by faster progression, and worse psychiatric and cognitive dysfunction, worsened by interventions such as DBS.^3, 5–8^ However, recent studies have contradicted these results, reporting similar cognitive and motor deterioration in *GBA1* variant carriers, regardless of variant type.^9-12^ Phase II trials are now underway evaluating changes in cognitive function at 12 months as primary outcomes, recruiting either mild or severe variant GBA-PD.^13^ An understanding of the natural clinical history of PD associated with different *GBA1* variants is crucial to appropriate trial design.

Here, we present our experience from a large, multi-centre, international case-control study incorporating PD patients negative (idiopathic PD-iPD) or positive for *GBA1* variants (GBA-PD), the latter group being representative of all variant types (risk, mild, and severe). We highlight how the severe *GBA1* variant carriers represent the only group with a remarkably different and more severe clinical phenotype compared to iPD. Our findings caution the PD community to avoid generalisation and consider GBA-PD as a clinically heterogeneous condition.

## Methods

### Study design and clinical data

Participants were recruited via the RAPSODI study (University College London) in the United Kingdom,^14^ and two neurology tertiary centres in Italy (IRCCS Mondino Foundation, Pavia, and Azienda USL-IRCCS, Reggio Emilia). *GBA1* variants were classified into four classes (severe, mild, risk, and unknown significance) as previously reported (a list of *GBA1* variants included in the study is presented in Supplementary Table 1).^15^ Information about family history, disease onset, and medications was collected, and levodopa equivalent daily dose (LEDD) calculated for each participant.^16, 17^ Motor and non-motor functions were evaluated through an extensive clinical examination (see Supplementary Methods).

This study was approved by the local Ethics Committees (London – Queen Square REC: 15/LO/1155; EC of Pavia: code P-20210009687; EC of Area Vasta Emilia Nord: code 2021/0092531). All participants signed informed consent upon enrolment.

## Statistical analysis

A first set of statistical analyses were completed comparing iPD to GBA-PD. In a second set, patients were stratified according to their *GBA1* variant severity into risk, mild, and severe (carriers of variants of unknown significance were excluded from this comparison because of the low numbers of individuals in this group). Study data were collected and managed using REDCap electronic data capture tools.^18^ Statistical analyses were performed using R version 4.2.3.^19^ Full statistical analysis methods can be found in Supplementary Methods.

## Results

Demographic and clinical features of iPD and GBA-PD groups are summarised in Table 1. A total of 311 PD patients (183 iPD, 128 GBA-PD) were recruited. Within the GBA-PD, 43% (N=55) carried *GBA1* risk variants, 30% (N=39) severe variants, 19% (N=24) mild variants, and 8% (N=10) carried variants of unknown significance.

**Table 1.**
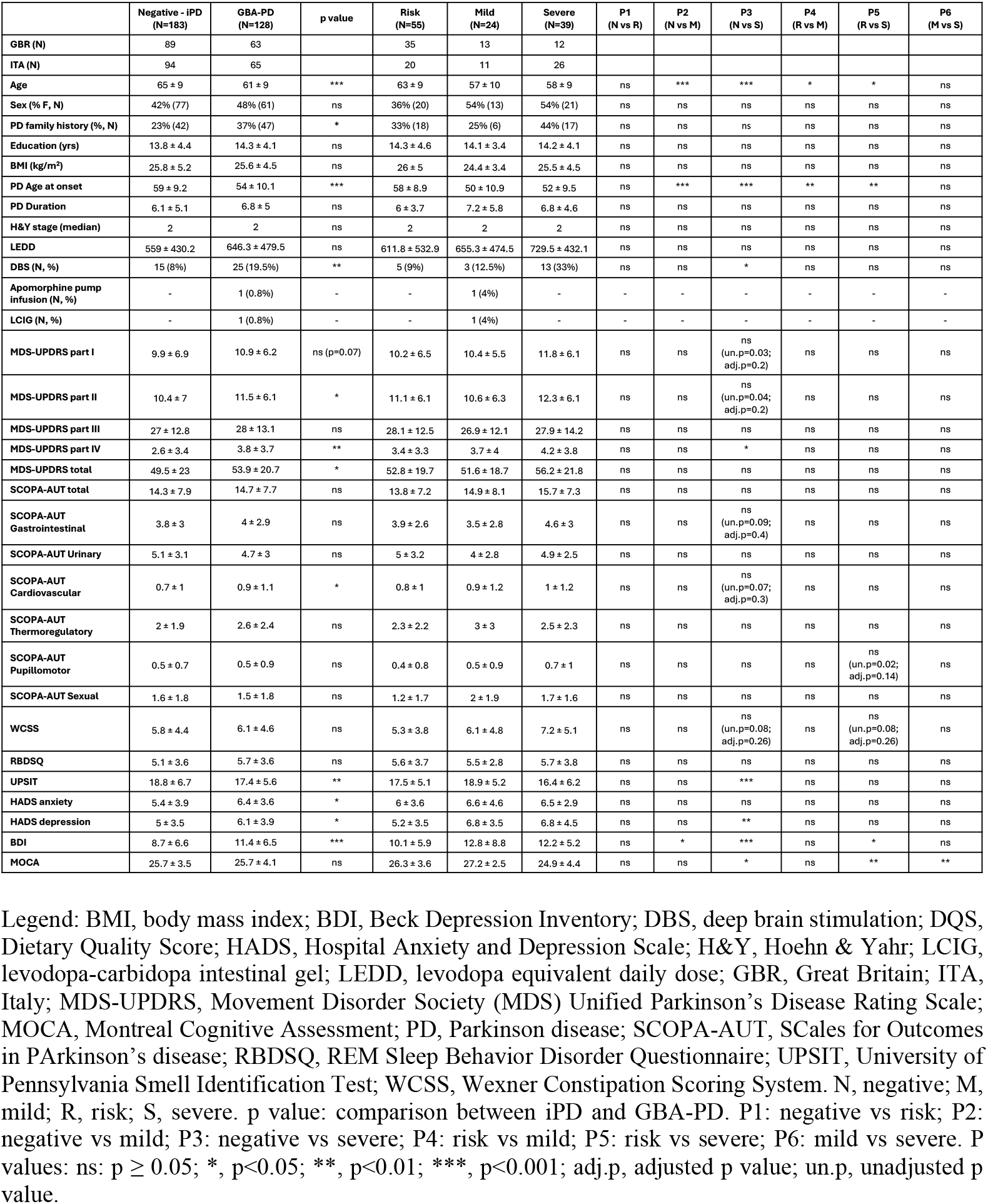
Demographics and clinical features of study cohort.

|  | Negative - iPD<br>(N=183) | GBA-PD<br>(N=128) | p value | Risk<br>(N=55) | Mild<br>(N=24) | Severe<br>(N=39) | P1<br>(N vs R) | P2<br>(N vs M) | P3<br>(N vs S) | P4<br>(R vs M) | P5<br>(R vs S) | P6<br>(M vs S) |
| --- | --- | --- | --- | --- | --- | --- | --- | --- | --- | --- | --- | --- |
| GBR (N) | 89 | 63 |  | 35 | 13 | 12 |  |  |  |  |  |  |
| ITA (N) | 94 | 65 |  | 20 | 11 | 26 |  |  |  |  |  |  |
| Age | 65 ± 9 | 61 ± 9 | *** | 63 ± 9 | 57 ± 10 | 58 ± 9 | ns | *** | *** | * | * | ns |
| Sex (% F, N) | 42% (77) | 48% (61) | ns | 36% (20) | 54% (13) | 54% (21) | ns | ns | ns | ns | ns | ns |
| PD family history (% N) | 23% (42) | 37% (47) | * | 33% (18) | 25% (6) | 44% (17) | ns | ns | ns | ns | ns | ns |
| Education (yrs) | 13.8 ± 4.4 | 14.3 ± 4.1 | ns | 14.3 ± 4.6 | 14.1 ± 3.4 | 14.2 ± 4.1 | ns | ns | ns | ns | ns | ns |
| BMI (kg/m<sup>2</sup>) | 25.8 ± 5.2 | 25.6 ± 4.5 | ns | 26 ± 5 | 24.4 ± 3.4 | 25.5 ± 4.5 | ns | ns | ns | ns | ns | ns |
| PD Age at onset | 59 ± 9.2 | 54 ± 10.1 | *** | 58 ± 8.9 | 50 ± 10.9 | 52 ± 9.5 | ns | *** | *** | ** | ** | ns |
| PD Duration | 6.1 ± 5.1 | 6.8 ± 5 | ns | 6 ± 3.7 | 7.2 ± 5.8 | 6.8 ± 4.6 | ns | ns | ns | ns | ns | ns |
| H&Y stage (median) | 2 | 2 | ns | 2 | 2 | 2 | ns | ns | ns | ns | ns | ns |
| LEDD | 559 ± 430.2 | 646.3 ± 479.5 | ns | 611.8 ± 532.9 | 655.3 ± 474.5 | 729.5 ± 432.1 | ns | ns | ns | ns | ns | ns |
| DBS (N, %) | 15 (8%) | 25 (19.5%) | ** | 5 (9%) | 3 (12.5%) | 13 (33%) | ns | ns | * | ns | ns | ns |
| Apomorphine pump infusion (N, %) | - | 1 (0.8%) | - | - | 1 (4%) | - | - | - | - | - | - | - |
| LCIG (N, %) | - | 1 (0.8%) | - | - | 1 (4%) | - | - | - | - | - | - | - |
| MDS-UPDRS part I | 9.9 ± 6.9 | 10.9 ± 6.2 | ns (p=0.07) | 10.2 ± 6.5 | 10.4 ± 5.5 | 11.8 ± 6.1 | ns | ns | ns<br>(un.p=0.03;<br>adj.p=0.2) | ns | ns | ns |
| MDS-UPDRS part II | 10.4 ± 7 | 11.5 ± 6.1 | * | 11.1 ± 6.1 | 10.6 ± 6.3 | 12.3 ± 6.1 | ns | ns | ns<br>(un.p=0.04;<br>adj.p=0.2) | ns | ns | ns |
| MDS-UPDRS part III | 27 ± 12.8 | 28 ± 13.1 | ns | 28.1 ± 12.5 | 26.9 ± 12.1 | 27.9 ± 14.2 | ns | ns | ns | ns | ns | ns |
| MDS-UPDRS part IV | 2.6 ± 3.4 | 3.8 ± 3.7 | ** | 3.4 ± 3.3 | 3.7 ± 4 | 4.2 ± 3.8 | ns | ns | * | ns | ns | ns |
| MDS-UPDRS total | 49.5 ± 23 | 53.9 ± 20.7 | * | 52.8 ± 19.7 | 51.6 ± 18.7 | 56.2 ± 21.8 | ns | ns | ns | ns | ns | ns |
| SCOPA-AUT total | 14.3 ± 7.9 | 14.7 ± 7.7 | ns | 13.8 ± 7.2 | 14.9 ± 8.1 | 15.7 ± 7.3 | ns | ns | ns | ns | ns | ns |
| SCOPA-AUT Gastrointestinal | 3.8 ± 3 | 4 ± 2.9 | ns | 3.9 ± 2.6 | 3.5 ± 2.8 | 4.6 ± 3 | ns | ns | ns<br>(un.p=0.09;<br>adj.p=0.4) | ns | ns | ns |
| SCOPA-AUT Urinary | 5.1 ± 3.1 | 4.7 ± 3 | ns | 5 ± 3.2 | 4 ± 2.8 | 4.9 ± 2.5 | ns | ns | ns | ns | ns | ns |
| SCOPA-AUT Cardiovascular | 0.7 ± 1 | 0.9 ± 1.1 | * | 0.8 ± 1 | 0.9 ± 1.2 | 1 ± 1.2 | ns | ns | ns<br>(un.p=0.07;<br>adj.p=0.3) | ns | ns | ns |
| SCOPA-AUT Thermoregulatory | 2 ± 1.9 | 2.6 ± 2.4 | ns | 2.3 ± 2.2 | 3 ± 3 | 2.5 ± 2.3 | ns | ns | ns | ns | ns | ns |
| SCOPA-AUT Pupillomotor | 0.5 ± 0.7 | 0.5 ± 0.9 | ns | 0.4 ± 0.8 | 0.5 ± 0.9 | 0.7 ± 1 | ns | ns | ns | ns | ns<br>(un.p=0.02;<br>adj.p=0.14) | ns |
| SCOPA-AUT Sexual | 1.6 ± 1.8 | 1.5 ± 1.8 | ns | 1.2 ± 1.7 | 2 ± 1.9 | 1.7 ± 1.6 | ns | ns | ns | ns | ns | ns |
| WCSS | 5.8 ± 4.4 | 6.1 ± 4.6 | ns | 5.3 ± 3.8 | 6.1 ± 4.8 | 7.2 ± 5.1 | ns | ns | ns<br>(un.p=0.08;<br>adj.p=0.26) | ns | ns<br>(un.p=0.08;<br>adj.p=0.26) | ns |
| RBDSQ | 5.1 ± 3.6 | 5.7 ± 3.6 | ns | 5.6 ± 3.7 | 5.5 ± 2.8 | 5.7 ± 3.8 | ns | ns | ns | ns | ns | ns |
| UPSIT | 18.8 ± 6.7 | 17.4 ± 5.6 | ** | 17.5 ± 5.1 | 18.9 ± 5.2 | 16.4 ± 6.2 | ns | ns | *** | ns | ns | ns |
| HADS anxiety | 5.4 ± 3.9 | 6.4 ± 3.6 | * | 6 ± 3.6 | 6.6 ± 4.6 | 6.5 ± 2.9 | ns | ns | ns | ns | ns | ns |
| HADS depression | 5 ± 3.5 | 6.1 ± 3.9 | * | 5.2 ± 3.5 | 6.8 ± 3.5 | 6.8 ± 4.5 | ns | ns | ** | ns | ns | ns |
| BDI | 8.7 ± 6.6 | 11.4 ± 6.5 | *** | 10.1 ± 5.9 | 12.8 ± 8.8 | 12.2 ± 5.2 | ns | * | *** | ns | * | ns |
| MOCA | 25.7 ± 3.5 | 25.7 ± 4.1 | ns | 26.3 ± 3.6 | 27.2 ± 2.5 | 24.9 ± 4.4 | ns | ns | * | ns | ** | ** |

Legend: BMI, body mass index; BDI, Beck Depression Inventory; DBS, deep brain stimulation; DQS, Dietary Quality Score; HADS, Hospital Anxiety and Depression Scale; H&Y, Hoehn & Yahr; LCIG, levodopa-carbidopa intestinal gel; LEDD, levodopa equivalent daily dose; GBR, Great Britain; ITA, Italy; MDS-UPDRS, Movement Disorder Society (MDS) Unified Parkinson's Disease Rating Scale; MOCA, Montreal Cognitive Assessment; PD, Parkinson disease; SCOPA-AUT, Scales for Outcomes in Parkinson's disease; RBDSQ, REM Sleep Behavior Disorder Questionnaire; UPSIT, University of Pennsylvania Smell Identification Test; WCSS, Wexner Constipation Scoring System. N, negative; M, mild; R, risk; S, severe. p value: comparison between iPD and GBA-PD. P1: negative vs risk; P2: negative vs mild; P3: negative vs severe; P4: risk vs mild; P5: risk vs severe; P6: mild vs severe. P values: ns, p ≥ 0.05; \*, p < 0.05; \*\*, p < 0.01; \*\*\*, p < 0.001; adj.p, adjusted p value; un.p, unadjusted p value.

### Two-group comparison: iPD vs GBA-PD

Compared with iPD, GBA-PD patients showed a significantly younger age (61 vs 65 years) and more frequent positive family history for PD (37% vs 23% of participants). No differences in sex distribution were found between groups. In terms of disease-associated features, GBA-PD patients developed the disease 5 years earlier and had undergone DBS more frequently than iPD patients. No differences in disease duration or LEDD were identified.

GBA-PD patients presented with more severe mood disorders (HADS depression: OR, 1.9; 95% CI: 1.2-3.1; p=0.01, and BDI: OR, 2.3; 95% CI: 1.5-3.7; p=0.0002), anxiety (HADS anxiety: OR, 1.8; 95% CI: 1.1-3; p=0.02), olfactory dysfunction (UPSIT: β=-2.2, p=0.002), subjective motor disability (MDS-UPDRS part II: p=0.03) and motor complications (MDS-UPDRS part IV: p=0.003), and greater cardiovascular dysfunction (p=0.03). Despite similar global cognitive function as measured by the total MOCA score, the GBA-PD group performed significantly worse in tasks evaluating visuospatial and executive functions (all p values<0.05, see Table 2; individual p values, ORs and CIs are reported in Supplementary Table 2).

**Table 2.** Performances in specific cognitive abilities (MOCA sub-scores).

| MOCA sub-scores | Negative – iPD (N=183) | GBA-PD (N=128) | p value | Risk (N=55) | Mild (N=24) | Severe (N=39) | P1 (N vs R) | P2 (N vs M) | P3 (N vs S) | P4 (R vs M) | P5 (R vs S) | P6 (M vs S) |
| --- | --- | --- | --- | --- | --- | --- | --- | --- | --- | --- | --- | --- |
| Diagram | 0.9 ± 0.4 | 0.8 ± 0.4 | * | 0.9 ± 0.4 | 0.8 ± 0.4 | 0.8 ± 0.4 | ns | * | ns (p=0.08) | ns | ns | ns |
| Cube | 0.7 ± 0.4 | 0.7 ± 0.5 | * | 0.8 ± 0.4 | 0.8 ± 0.4 | 0.6 ± 0.5 | ns | ns | *** | ns | * | ns |
| Clock | 2.6 ± 0.7 | 2.5 ± 0.8 | * | 2.5 ± 0.8 | 2.6 ± 0.6 | 2.4 ± 0.8 | ns | ns | * | ns | ns | ns |
| Visuospatial/executive functions | 4.2 ± 1.2 | 4 ± 1.4 | * | 4.1 ± 1.3 | 4.2 ± 1.2 | 3.8 ± 1.4 | ns | ns | ** | ns | * | ns |
| Naming | 3 ± 0.3 | 3 ± 0.5 | ns | 3 ± 0.4 | 3 ± 0.3 | 3 ± 0.6 | ns | ns | ns | ns | ns | ns |
| Digits | 1.8 ± 0.5 | 1.8 ± 0.4 | ns | 1.8 ± 0.4 | 1.9 ± 0.3 | 1.8 ± 0.5 | ns | ns | ns | ns | ns | ns |
| Letters | 0.9 ± 0.3 | 0.9 ± 0.3 | ns | 1 ± 0.2 | 1 ± 0.2 | 0.8 ± 0.4 | ns | ns | ns | ns | * | ns |
| Subtraction | 2.7 ± 0.6 | 2.7 ± 0.7 | ns | 2.8 ± 0.5 | 2.9 ± 0.3 | 2.4 ± 0.9 | ns | ns | ns (p=0.08) | ns | * | ns (p=0.054) |
| Attention | 5.4 ± 0.9 | 5.4 ± 1.1 | ns | 5.6 ± 0.7 | 5.7 ± 0.6 | 5.1 ± 1.3 | ns | ns | ns | ns | * | * |
| Repeat | 1.7 ± 0.5 | 1.8 ± 0.5 | ns | 1.7 ± 0.5 | 1.8 ± 0.4 | 1.8 ± 0.4 | ns | ns | ns | ns | ns | ns |
| Fluency | 0.8 ± 0.4 | 0.8 ± 0.4 | ns | 0.7 ± 0.4 | 0.9 ± 0.4 | 0.7 ± 0.4 | ns | ns | ns | ns | ns | ns |
| Language | 2.5 ± 0.7 | 2.5 ± 0.7 | ns | 2.5 ± 0.7 | 2.7 ± 0.6 | 2.5 ± 0.7 | ns | ns | ns | ns | ns | ns |
| Abstraction | 1.7 ± 0.5 | 1.7 ± 0.5 | ns | 1.8 ± 0.5 | 1.8 ± 0.4 | 1.6 ± 0.6 | ns | ns | ns | ns | ns | ns |
| Delayed recall | 2.8 ± 1.7 | 3.2 ± 1.5 | ns | 3.3 ± 1.5 | 3.8 ± 1.2 | 3 ± 1.5 | ns | ns | ns | ns | ns | ns |
| Orientation | 5.9 ± 0.4 | 5.8 ± 0.6 | ns | 5.9 ± 0.5 | 5.9 ± 0.3 | 5.7 ± 0.7 | ns | ns | ns | ns | ns | ns |

Legend: PD, Parkinson disease. N, negative; M, mild; R, risk; S, severe. p value: comparison between iPD and GBA-PD. P1: negative vs risk; P2: negative vs mild; P3: negative vs severe; P4: risk vs mild; P5: risk vs severe; P6: mild vs severe. P values: ns: $p \geq 0.05$ ; \*, $p < 0.05$ ; \*\*, $p < 0.01$ ; \*\*\*, $p < 0.001$ .

### Clinical features in stratified *GBA1*-variant type groups

When patients were stratified according to *GBA1* variant type, severe and mild GBA-PD were significantly younger than both risk GBA-PD (p=0.02, and p=0.01, respectively) and iPD (both p<0.001), with no difference in age between severe and mild GBA-PD. Severe and mild GBA-PD also developed PD earlier than risk GBA-PD (p=0.006 and p=0.003, respectively) and iPD (both p<0.001). Severe GBA-PD patients had a more frequent positive family history for PD (44%) and were more frequently subjected to DBS procedures compared to iPD patients (Table 1). Groups were homogenous in terms of disease duration and LEDD.

When clinical features were compared, severe GBA-PD patients presented with a distinct clinical profile when compared to iPD patients, but not to the risk or mild GBA-PD patients (Table 1). This profile was characterised by more severe depression (HADS depression: OR, 2.7; 95% CI: 1.3- 5.3; p=0.005, and BDI: OR, 3.8; 95% CI: 2-7.4; p<0.0001), olfactory dysfunction (UPSIT: β=-3.8, p=0.0006), cognitive impairment (MOCA: β=-1.5, p=0.01), motor complications (p.adj=0.04) and a trend for more severe constipation/gastrointestinal dysfunction (see Table 1). Cognitive performances were worse in severe GBA-PD, also compared with risk and mild GBA-PD patients (all p values<0.01); these results remained statistically significant even after removing subjects with DBS, with only a nearly significance for the comparison between mild and severe carriers (p=0.0505), which might be due to low numbers of individuals (21 and 26, respectively) and thus loss of statistical power. Severe GBA-PD were mostly affected in visuospatial and executive function and attention abilities (Table 2 and Supplementary Table 2). No other statistically significant differences emerged among the four groups in the severity of motor and non-motor symptoms when results were adjusted for multiple comparisons.

## Discussion

In this large, multi-centre, international case-control study, we demonstrated the importance of considering the type of *GBA1* genetic variant in research studies and clinical trials conducted on GBA-PD. Reporting findings on GBA-PD without stratifying according to *GBA1* variant type, can be misleading: differences can be detected related to iPD, however these are mainly driven by severe variant carriers.

Our results align with previous findings showing a more severe phenotype in severe variant carriers.^6, 8, 20^ In a previous study comparing a large cohort of GBA-PD (139 mild, 48 severe) to 152 iPD patients, more severe depression, hallucinations, worse hyposmia, and higher frequencies of REM sleep behaviour disorder (RBD) were detected in severe GBA-PD compared to the other two groups.^20^ However, this study did not include risk variant carriers, who numerically represent the predominant group of GBA-PD individuals, especially in those of Caucasian background.^1^

In contrast to previous reports, we did not detect any significant cognitive impairment in carriers of risk or mild variants. One study found that both pathogenic (mix of mild and severe) variant carriers (N=60) and p.E365K carriers (N=65) had higher incidence of dementia and a greater impairment in working memory/executive function and visuospatial abilities compared to iPD, however the disease duration in these groups was mostly >7.5years, so additional factors beyond *GBA1* status might have contributed to these results.^11^ Cognitive decline at 7 years from diagnosis was faster in pathogenic and risk variant carriers than iPD in another study, however, the association with progression rate to dementia was much smaller in the risk variant carriers group, and no distinction was made between severe and mild variant carriers.^21^

Our results showed more severe cardiovascular dysfunction in GBA-PD vs iPD, but we did not see any differences across variant types. Previous reports also found more impaired cardiovascular autonomic control characterised by a lower parasympathetic modulation at rest and a lower parasympathetic modulation in response to active standing, in GBA-PD compared to iPD.^22^ GBA-PD also presented a lower heart-to-mediastinum uptake ratio in ^123^I-meta-iodobenzylguanidine scintigraphy suggestive of reduced cardiac sympathetic denervation,^23^ and greater cardiac sympatho-vagal demodulation.^24^ The latter study detected greater impairment in cardiovagal function in severe GBA-PD compared to risk and mild GBA-PD combined together, but numbers were small.^24^ Nonetheless, these findings make this area worth investigating in the future. In our study, we observed higher scores in the SCOPA pupillomotor subdomain in severe GBA-PD, which might be related to a more extreme pupillary parasympathetic dysfunction in this group. Interestingly, pupillomotor abnormalities suggestive of dysfunctional parasympathetic innervation were also detected in individuals with neuronopathic Gaucher disease, caused by biallelic pathogenic variants in *GBA1*,^25^ or with Fabry disease, another lysosomal storage disorder.^26^

Several disease-modifying therapies targeting the *GBA1* pathway are in Phase II/III of clinical trial stage, aiming to halt/slow GBA-PD progression.^4, 27^ Recently, a Phase II, randomized-controlled trial (MOVES-PD), testing the effect of the glucosylceramide synthase inhibitor Venglustat in GBA-PD patients, did not meet its primary endpoint, and showed paradoxical worsening of motor symptoms in mild GBA-PD treated with Venglustat.^28^ This highlight the need to refine patients’ selection and stratify individuals for variant severity in trials targeting the GBA-PD population in order to optimise chances of success.^28^

Our study has some limitations. First, the relatively small number of individuals with severe and mild GBA-PD might have undermined the results. Second, this is a case-control study, however a 2-year follow-up assessment of this cohort is in progress.

Despite these limitations, we believe that this study provides clinicians with useful information to approach their GBA-PD patients in the clinic, tailoring their counselling on prognosis and disease trajectories, and integrating our recent consensus guidance on PD risk.^1^ Moreover, we hope that these findings will inform researchers involved in clinical trials to improve the design of such studies and guide the choice of realistic outcomes in the correct patient groups. Finally, this study provides insight into the pathophysiological mechanisms underlying disease severity in GBA-PD.

## Data Availability

All data produced in the present study are available upon reasonable request to the authors.

## Acknowledgments

This research was funded in part by Aligning Science Across Parkinson’s (Grant number: ASAP- 000420) through the Michael J. Fox Foundation for Parkinson’s Research (MJFF) and by the EU Joint Programme—Neurodegenerative Research (JPND: GBA-PaCTS) through the MRC grant code MR/T046007/1. For the purpose of open access, the author has applied a CC BY 4.0 public copyright license to all Author Accepted Manuscripts arising from this submission.

## Authors’ Roles

1. Research project: A. Conception, B. Organization, C. Execution.
2. Statistical Analysis: A. Design, B. Execution, C. Review and Critique.
3. Manuscript Preparation: A. Writing of the first draft, B. Review and Critique.

EM: 1A, 1B, 1C, 2A, 2B, 2C, 3A.

SLDP: 1A, 1B, 1C, 3B.

JM: 1A, 1B, 1C, 3B.

RM: 1B, 1C, 3B.

SK: 1B, 3B.

PM: 1B, 1C, 3B.

LG: 1B, 1C, 3B.

RC: 1B, 1C, 3B.

MT: 1B, 1C, 3B.

NL: 1B, 1C, 3B.

FV: 1C, 3B.

FC: 1C, 3B.

VF: 1C, 3B.

SY: 1B, 1C, 3B.

NL: 1B, 1C, 3B.

FB: 1A, 1B, 2C, 3B.

MA: 1A, 1B, 1C, 2C, 3B.

AHVS: 1A, 1B, 2C, 3B.

