## Supplementary Material for "Severe *GBA1* variants drive the GBA-PD clinical phenotype: implications for counselling and clinical trials"

**Table of Contents:**

- **Supplementary Methods (p. 2-3)**
- **Supplementary Tables (p. 4-5)**
- **References (p. 6)**

**Supplementary Methods**

**Recruitment strategy and *GBA1* sequencing**

Participants were recruited from the United Kingdom and Italy. Prior to enrolment, participants were screened for variants in the *GBA1* gene.

For the UK cohort, participants were recruited via the RAPSODI GD and PD Frontline portals, from University College London (UCL).^1^ Full *GBA1* gene sequencing was performed on saliva samples collected with the DNA OG-500 kit from DNA Genotek and posted back from participants. DNA extraction and long range sequencing of an 8.9 kb amplicon, including all coding exons and introns of *GBA1*, was performed using bespoke primers on the Oxford Nanopore MinION platform, as previously described.^2^ This was done partially at the laboratories of the Department of Clinical and Movement Neurosciences, UCL Institute of Neurology, United Kingdom, and partially at the Exeter Clinical Laboratory International, United Kingdom, a diagnostic clinical laboratory accredited by the UK’s National Accreditation Body (UKAS 8092). *GBA1* positive results were confirmed by Sanger sequencing at the Exeter Clinical Laboratory International. Copy number variants (both reciprocal duplication alleles and reciprocal fusion alleles) were evaluated on saliva samples using Oxford Nanopore sequencing after polymerase chain reaction (PCR) enrichment at the laboratories of the Department of Clinical and Movement Neurosciences, UCL Institute of Neurology, as previously described.^3^ The UK participants were also screened for the *LRRK2* G2019S variant, as an exclusion criterion. The *LRRK2* gene was genotyped with KBiosciences Competitive AlleleSpecific PCR SNP genotyping system at the LGC Genomics, Hoddesdon, Herts.

For the Italian cohort, participants were recruited from two neurology tertiary centres, the IRCCS Mondino Foundation in Pavia, and the Neurology Unit, Neuromotor & Rehabilitation Department, Azienda USL-IRCCS in Reggio Emilia. Analysis of the *GBA1* gene in the Mondino Pavia cohort was performed by a next-generation sequencing (NGS)-based method, which relies on the selective amplification of the whole *GBA1* gene in one long PCR fragment (6kb) followed by Nextera sequencing and a customized bioinformatics pipeline aimed at masking the *GBAP1* pseudogene. Identified variants were validated by conventional Sanger sequencing through *GBA1* amplification in three overlapping fragments using specific primer pairs. Pathogenic variants in 15 PD-related genes associated with autosomal dominant (*SNCA*, *LRRK2*, *VPS35*, *GBA1*), X-linked (*RAB39B*), and autosomal recessive PD (*PRKN*, *PINK1*, *PARK7*, *ATP13A2*, *PLA2G6*, *DNAJC6*, *SYNJ1*, *FBXO7*, *VPS13C*, *PTRHD1*) were searched for by means of NGS-based sequencing of Parkinson disease (PD) gene virtual panel as well as MLPA analysis (SALSA Kit P51-P52, MRC Holland). In the Reggio Emilia cohort, analysis of the *GBA1* gene for PD patients was performed by combining a primary *GBA1*-specific long-range PCR with subsequent *GBA1* exon-specific PCR and NGS of the resulting products, as previously described.^4^ Patients were also tested for 11 pathogenic or likely pathogenic *LRKK2* variants. If negative, 50 target genes were analysed by means of NGS-based sequencing of PD gene virtual panel, as previously described.^4^ For non-parkinsonian individuals of the Reggio Emilia cohort, the genetic analysis of the *GBA1* gene was performed either by long PCR approach or NGS-based method, as previously described.^5,6^

**Clinical evaluation**

Demographic information was collected from each participant enrolled in the study. An extensive range of clinical scales and questionnaires was administered to each participant, to evaluate motor and non-motor (autonomic function, constipation, rapid eye movements-REM-sleep behaviour disorder, olfaction, mood, cognition) symptoms of PD. These included: Movement Disorder Society-Unified Parkinson’s Disease Rating Scale MDS-UPDRS (part I-IV),^7^ Hoehn & Yahr (H&Y) scale,^8^ SCales for Outcomes in PArkinson’s disease (SCOPA-AUT),^9^ Wexner Constipation Scoring System (WCSS),^10^ REM Sleep Behaviour Disorder Questionnaire (RBDSQ),^11^ University of Pennsylvania Smell Identification Test (UPSIT),^12^ Hospital Anxiety and Depression Scale (HADS),^13^ Beck Depression Inventory (BDI),^14^ and Montreal Cognitive Assessment (MOCA).^15^

**Statistical analysis**

Categorical variables were compared using χ2 test. Continuous variables are presented as means ± SDs, unless otherwise specified. Group demographics and clinical variables were compared using t-test or non-parametric Wilcoxon rank-sum test for two-group comparison (iPD vs GBA-PD), or One-way ANOVA or non-parametric Kruskal-Wallis test for multiple-group comparisons. Multiple pairwise t-tests or the non-parametric analogue Dunn’s pairwise tests, with the Benjamini-Hochberg (BH) false discovery rate, were applied to correct for multiple comparison, with a significant threshold of 0.05. For HADS, BDI, and RBDSQ scores, data were categorised using clinically validated thresholds and ordinal logistic regression (OLR) was used to determine associations between categorised variables and the grouping variable, adjusted for covariates. For UPSIT and MOCA scores, a linear regression model was applied with adjustment for covariates. Sub-scores of the MOCA scale were analysed using ORL or binary regression according to variable type.

**Supplementary Table 1. List of *GBA1* variants.**

| **Genotype** | **N** | **Variant severity** |
| --- | --- | --- |
| G241R/WT | 2 | severe |
| H294Q+D448H/WT | 1 | severe |
| IVS2+1G>A/WT | 1 | severe |
| K(-27)R+L483P+P491P/WT | 1 | severe |
| L483P/WT | 18 | severe |
| N227S/WT | 3 | severe |
| P284T/WT | 1 | severe |
| Q239Ter/WT | 1 | severe |
| Q401Ter/WT | 1 | severe |
| R170C/WT | 1 | severe |
| R202Ter/WT | 1 | severe |
| R296Q/WT | 1 | severe |
| R296Ter/WT | 1 | severe |
| R502C/WT | 4 | severe |
| RecNcil/WT | 1 | severe |
| V433L/WT | 1 | severe |
| N409S/WT | 22 | mild |
| R209P/WT | 1 | mild |
| R368C/WT | 1 | mild |
| E365K/WT | 33 | risk |
| T408M/E365K | 2 | risk |
| T408M/WT | 20 | risk |
| E365D/WT | 1 | unknown |
| M124V/WT | 1 | unknown |
| M319T/WT | 1 | unknown |
| R301H/WT | 1 | unknown |
| R78H/WT | 1 | unknown |
| S149P/WT | 1 | unknown |
| S212T/WT | 1 | unknown |
| T125P/WT | 1 | unknown |
| T247S/WT | 1 | unknown |
| T306I/WT | 1 | unknown |

**Supplementary Table 2. Summary of statistical analyses for significant comparisons between groups in MOCA sub-scores.**

| **MOCA**  **sub-scores** | **iPD vs GBA-PD** | | | ***N vs M*** | | | ***N vs S*** | | | ***R vs S*** | | | ***M vs S*** | | |
| --- | --- | --- | --- | --- | --- | --- | --- | --- | --- | --- | --- | --- | --- | --- | --- |
|  | ***p*** | **OR** | **95% CI** | ***p*** | **OR** | **95% CI** | ***p*** | **OR** | **95% CI** | ***p*** | **OR** | **95% CI** | ***p*** | **OR** | **95% CI** |
| **Diagram** | 0.02 | 0.4 | 0.2-0.9 | 0.04 | 0.3 | 0.08-0.99 | ns | ns | ns | ns | ns | ns | ns | ns | ns |
| **Cube** | 0.02 | 0.5 | 0.3-0.9 | ns | ns | ns | 0.0009 | 0.2 | 0.09-0.5 | 0.01 | 0.3 | 0.09-0.7 | ns | ns | ns |
| **Clock** | 0.04 | 0.6 | 0.3-0.9 | ns | ns | ns | 0.02 | 0.4 | 0.2-0.9 | ns | ns | ns | ns | ns | ns |
| **Visuospatial/**  **executive f.** | 0.01 | 0.5 | 0.3-0.9 | ns | ns | ns | 0.002 | 0.3 | 0.2-0.7 | 0.02 | 0.4 | 0.2-0.9 | ns | ns | ns |
| **Letters** | ns | ns | ns | ns | ns | ns | ns | ns | ns | 0.04 | 0.2 | 0.02-0.8 | ns | ns | ns |
| **Subtraction** | ns | ns | ns | ns | ns | ns | ns | ns | ns | 0.01 | 0.3 | 0.09-0.8 | ns | ns | ns |
| **Attention** | ns | ns | ns | ns | ns | ns | ns | ns | ns | 0.01 | 0.3 | 0.1-0.8 | 0.03 | 0.3 | 0.09-0.9 |

Legend: CI, confidence interval; OR, odds ratio; PD, Parkinson disease; N, negative; M, mild; R, risk; S, severe.
